# Public acceptability of non-pharmaceutical interventions to control a pandemic in the United Kingdom: a discrete choice experiment

**DOI:** 10.1101/2021.10.12.21264883

**Authors:** Luis Enrique Loria-Rebolledo, Mandy Ryan, Verity Watson, Mesfin G Genie, Ruben Andreas Sakowsky, Daniel Powell, Shantini Paranjothy

**Affiliations:** Health Economics Research Unit, University of Aberdeen, UK AB25 2ZD; Department of Medical Ethics and History of Medicine, University Medical Center Göttingen, Germany; Health Data Science Research Centre, University of Aberdeen, UK, AB25 2ZD; Health Psychology, University of Aberdeen, UK, AB25 2ZD

## Abstract

**Objective:** To understand how individuals make trade-offs between features of lockdown interventions to control a pandemic across the four nations of the United Kingdom.

**Design:** Survey that included a Discrete Choice Experiment (DCE). The survey design was informed using policy documents, social media analysis and with input from remote think aloud interviews with members of the public (n=23).

**Setting:** Nation-wide survey across the four nations of the United Kingdom. Representative sample in terms of age and sex for each of the nations recruited using an online panel between 29^th^ October and 12^th^ December 2020.

**Participants:** Individuals who are over 18 years old. A total of 4120 adults completed the survey (1112 in England, 848 in Northern Ireland, 1143 in Scotland and 1098 in Wales).

**Primary outcome measure:** Adult’s preferences for, and trade-offs between, type of lockdown restrictions, length of lockdown, postponement of routine healthcare, excess deaths, impact on ability to buy things and unemployment.

**Results:** In all four countries, one out of five respondents were willing to reduce excess deaths at all costs. The majority of adults are willing to accept higher excess deaths if this means lockdowns that are less strict, shorter and do not postpone routine healthcare. On average, respondents in England were willing to accept a higher increase in excess deaths to have less strict lockdown restrictions introduced compared to Scotland, Northern Ireland, and Wales, respectively.

**Conclusions:** The majority of the UK population is willing to accept the increase in excess deaths associated with introducing less strict lockdown restrictions. The acceptability of different restriction scenarios varies according to the features of the lockdown and across countries. Authorities can use information about trade-off preferences to inform the introduction of different lockdown restriction levels, and design compensation policies that maximise societal welfare.

**Strengths and limitations of this study:**

- This study offers empirical evidence that, unlike existing data from opinion polls and citizens’ panels, offers a clear understanding of the trade-offs between restrictions and impacts of lockdown on society.
- Estimating preferences for each nation, and quantifying them in terms of a common denominator, allows a comparison that takes into account the heterogeneity of UK nations and can be used to inform the introduction of different levels of lockdown restrictions in each.
- A limitation of our study is that we are not able to estimate the effect of on-going lockdowns in preferences. Furthermore, our results are not necessarily transferable to other nations.

## Introduction

The COVID-19 pandemic has required countries worldwide to introduce non-pharmaceutical interventions to protect the health and wellbeing of their citizens.[1] The majority of European and high-income nations have focused on reducing the R number to less than one and thereby curtailing the epidemic spread of the virus.[2,3] This strategy requires a number of non-pharmaceutical interventions such as enforced social distancing across all age groups, closing schools and non-essential businesses, and a range of other social restrictions.[4] This has led to local and nationwide lockdowns and other restrictions to control infection rates and excess deaths within geographically defined populations.[5-7]

Lockdowns have wider indirect impacts on health and wellbeing, and lockdown decisions require a careful balancing of the direct impacts on mortality caused by COVID-19 with the indirect wider health, social and economic impacts.[8-11] Further, lockdown compliance will determine its effectiveness. Compliance is more likely if policies are acceptable. Policies are more likely to be acceptable if the public’s preferences are understood and the diversity of view is recognised. The World Health Organization criteria for deciding whether to lift lockdown restrictions is defined as *“Communities are fully educated, engaged and empowered to adjust to the “new norm” of everyday life*.[12] This criterion requires a better understanding of how the public respond to and value the trade-offs faced during and post-pandemic. For example, are the public willing to accept a certain number of excess deaths to have restrictions eased?

Prior to the COVID-19 pandemic, there was limited evidence on the understanding of how people think of lockdown policies in the UK.[13] During the pandemic, public attitudes to government responses to the pandemic have been explored using opinion polls and qualitative studies.[14-16] The Scottish Government and Bank of England established citizen’s panels.[17,18] These instruments offer insight into the views and concerns of the population. However, they provide no understanding of the trade-offs that individuals are willing to make. For example, the Scottish citizen’s panel recommended that the Scottish Government should implement an elimination strategy, and where this is not feasible, should aim for maximum suppression of the virus, but not the cost of the restrictions that were acceptable to achieve this. Thus, we use a preference elicitation instrument tailored to quantify preferences, a discrete choice experiment (DCE), to provide new evidence on the acceptable number of excess deaths to the UK public when easing or tightening restrictions.

## Methods

### Study Sample

We conducted a cross-sectional survey among a representative sample of adults aged over 18 from across the four nations of the United Kingdom. The survey was implemented between 29^th^ October and 12^th^ December 2020. Respondents were recruited using an online survey research panel maintained by the company Qualtrics. The survey was piloted in early October 2020 (n=50 per nation). Respondents were screened by the recruiting company using sex and age using quotas to ensure a balance in each nation. The research company excluded respondents that completed the survey in less than half the median time of completion of the pilot stage of the survey (14 minutes).

### Discrete Choice Experiment

Respondents completed a self-complete online survey that asked about the individual’s experience during the COVID-19 pandemic, lockdowns that had occurred, any impacts on their healthcare, their spending ability and employment. The survey included a discrete choice experiment (DCE), a choice-based survey that quantifies preferences for attributes (or features) of goods, services or policies. Respondents completed a series of eight choice tasks based on the features of government restrictions. The hypothetical choice tasks focussed on six features of government restrictions that describe different types of lockdown and their likely health and economic consequences.

Features used to describe the *type of lockdown* were: restriction severity using a colour-based tier system (Figure 1), length in weeks, and postponement of routine healthcare procedures. The *health consequences* were the number of excess deaths (we also report infection numbers as a complement based on the infection rate).[19] The *economic consequences* included respondent’s household’s ability to buy things (personal impact) and the number of job losses (societal impact). See the online Supplemental Table 1 for the features and associated levels. The features and levels were informed by policy documents,[12] impacts of interventions that were implemented in response to COVID-19,[4] literature on preferences for lockdown measures from previous pandemics,[20,21] and a social media analysis. A more detailed description of the development stage can be found in the study’s published protocol.[22]

**Figure 1:**
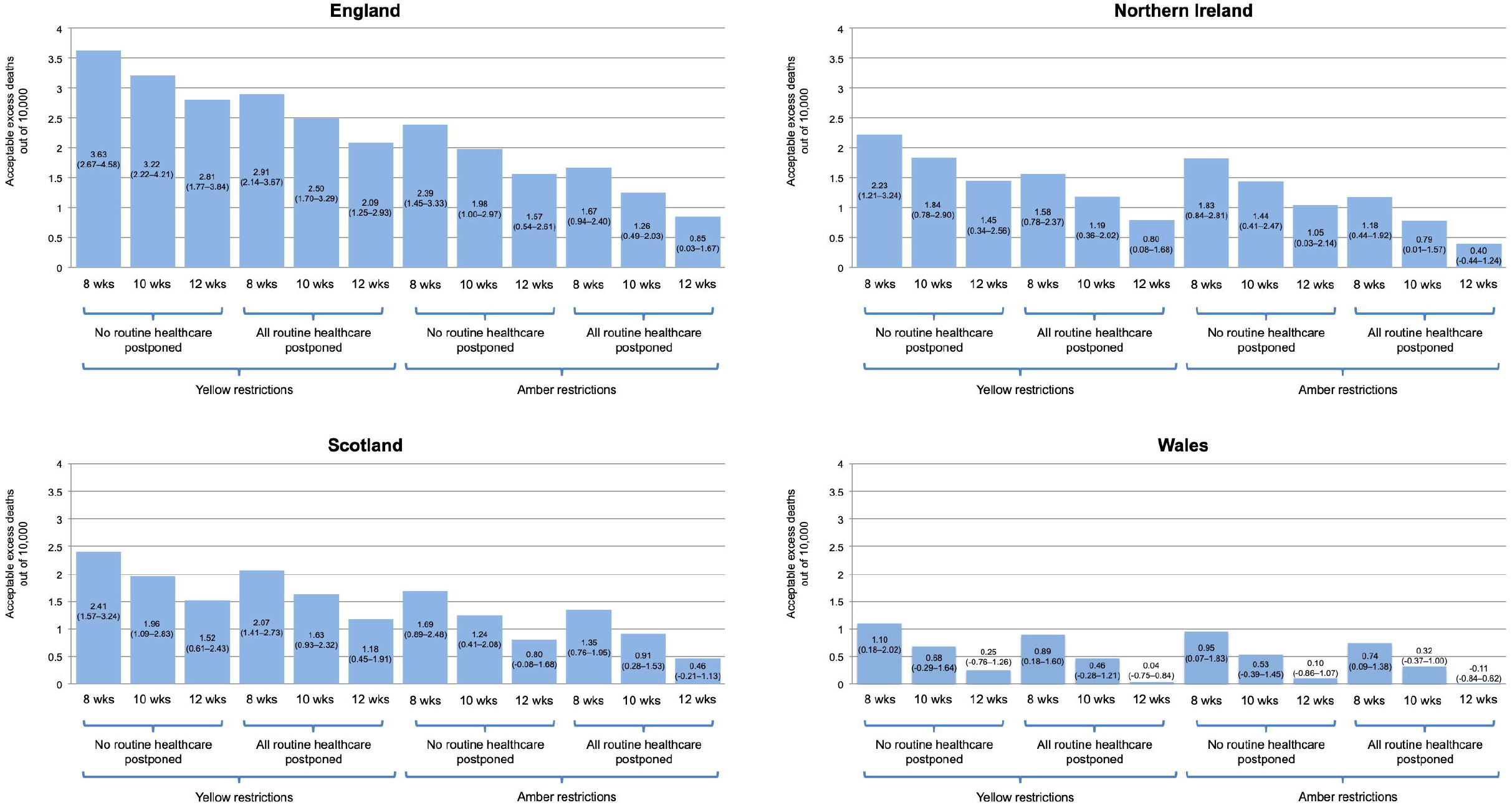

**Table 1.**
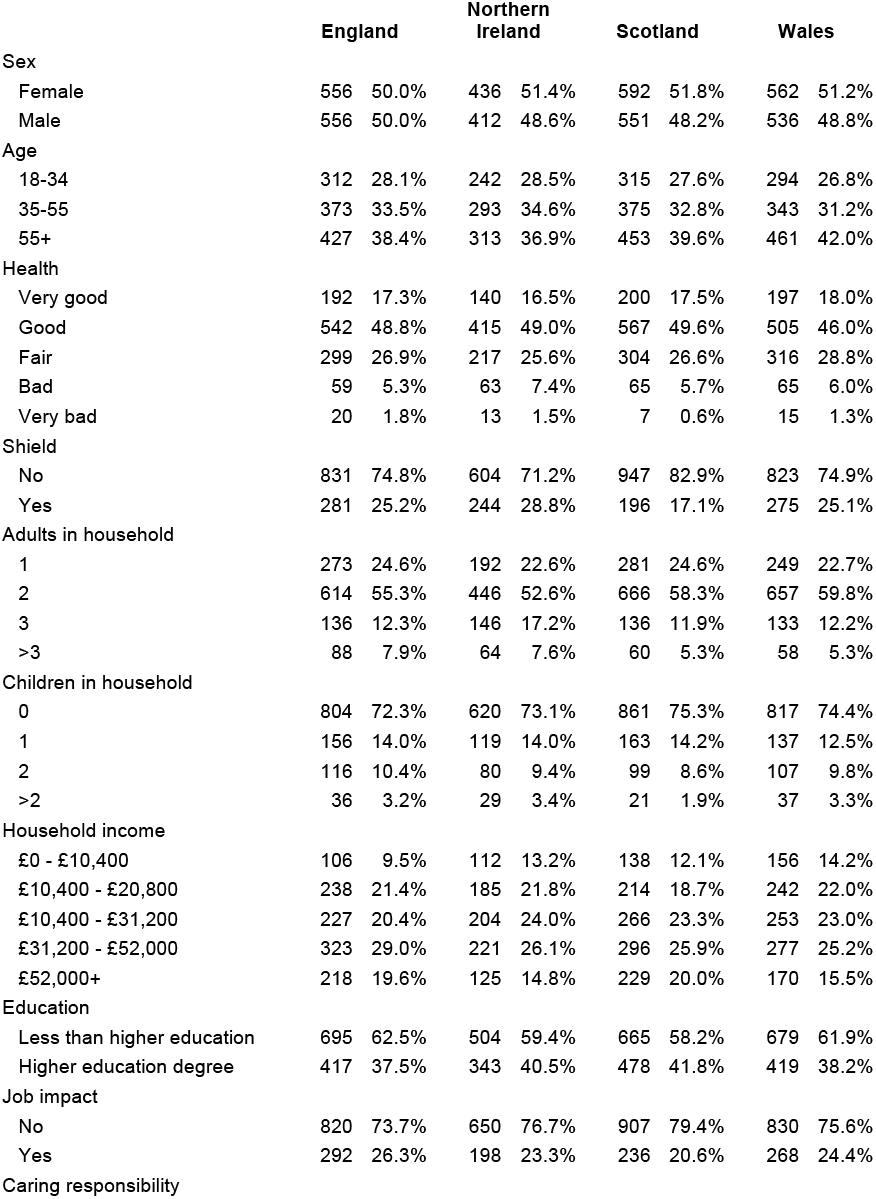
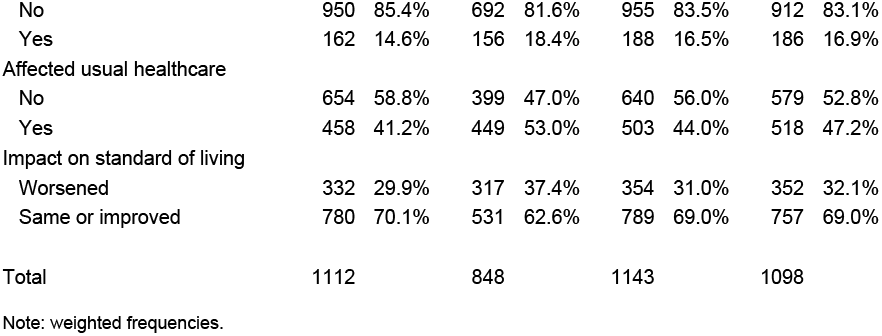
Characteristics associated with sample by nation.

|  | **England** |  | **Northern**<br>**Ireland** |  | **Scotland** |  | **Wales** |  |
| --- | --- | --- | --- | --- | --- | --- | --- | --- |
| Sex |  |  |  |  |  |  |  |  |
| Female | 556 | 50.0% | 436 | 51.4% | 592 | 51.8% | 562 | 51.2% |
| Male | 556 | 50.0% | 412 | 48.6% | 551 | 48.2% | 536 | 48.8% |
| Age |  |  |  |  |  |  |  |  |
| 18-34 | 312 | 28.1% | 242 | 28.5% | 315 | 27.6% | 294 | 26.8% |
| 35-55 | 373 | 33.5% | 293 | 34.6% | 375 | 32.8% | 343 | 31.2% |
| 55+ | 427 | 38.4% | 313 | 36.9% | 453 | 39.6% | 461 | 42.0% |
| Health |  |  |  |  |  |  |  |  |
| Very good | 192 | 17.3% | 140 | 16.5% | 200 | 17.5% | 197 | 18.0% |
| Good | 542 | 48.8% | 415 | 49.0% | 567 | 49.6% | 505 | 46.0% |
| Fair | 299 | 26.9% | 217 | 25.6% | 304 | 26.6% | 316 | 28.8% |
| Bad | 59 | 5.3% | 63 | 7.4% | 65 | 5.7% | 65 | 6.0% |
| Very bad | 20 | 1.8% | 13 | 1.5% | 7 | 0.6% | 15 | 1.3% |
| Shield |  |  |  |  |  |  |  |  |
| No | 831 | 74.8% | 604 | 71.2% | 947 | 82.9% | 823 | 74.9% |
| Yes | 281 | 25.2% | 244 | 28.8% | 196 | 17.1% | 275 | 25.1% |
| Adults in household |  |  |  |  |  |  |  |  |
| 1 | 273 | 24.6% | 192 | 22.6% | 281 | 24.6% | 249 | 22.7% |
| 2 | 614 | 55.3% | 446 | 52.6% | 666 | 58.3% | 657 | 59.8% |
| 3 | 136 | 12.3% | 146 | 17.2% | 136 | 11.9% | 133 | 12.2% |
| >3 | 88 | 7.9% | 64 | 7.6% | 60 | 5.3% | 58 | 5.3% |
| Children in household |  |  |  |  |  |  |  |  |
| 0 | 804 | 72.3% | 620 | 73.1% | 861 | 75.3% | 817 | 74.4% |
| 1 | 156 | 14.0% | 119 | 14.0% | 163 | 14.2% | 137 | 12.5% |
| 2 | 116 | 10.4% | 80 | 9.4% | 99 | 8.6% | 107 | 9.8% |
| >2 | 36 | 3.2% | 29 | 3.4% | 21 | 1.9% | 37 | 3.3% |
| Household income |  |  |  |  |  |  |  |  |
| £0 - £10,400 | 106 | 9.5% | 112 | 13.2% | 138 | 12.1% | 156 | 14.2% |
| £10,400 - £20,800 | 238 | 21.4% | 185 | 21.8% | 214 | 18.7% | 242 | 22.0% |
| £10,400 - £31,200 | 227 | 20.4% | 204 | 24.0% | 266 | 23.3% | 253 | 23.0% |
| £31,200 - £52,000 | 323 | 29.0% | 221 | 26.1% | 296 | 25.9% | 277 | 25.2% |
| £52,000+ | 218 | 19.6% | 125 | 14.8% | 229 | 20.0% | 170 | 15.5% |
| Education |  |  |  |  |  |  |  |  |
| Less than higher education | 695 | 62.5% | 504 | 59.4% | 665 | 58.2% | 679 | 61.9% |
| Higher education degree | 417 | 37.5% | 343 | 40.5% | 478 | 41.8% | 419 | 38.2% |
| Job impact |  |  |  |  |  |  |  |  |
| No | 820 | 73.7% | 650 | 76.7% | 907 | 79.4% | 830 | 75.6% |
| Yes | 292 | 26.3% | 198 | 23.3% | 236 | 20.6% | 268 | 24.4% |
| Caring responsibility |  |  |  |  |  |  |  |  |
| No | 950 | 85.4% | 692 | 81.6% | 955 | 83.5% | 912 | 83.1% |
| Yes | 162 | 14.6% | 156 | 18.4% | 188 | 16.5% | 186 | 16.9% |
| Affected usual healthcare |  |  |  |  |  |  |  |  |
| No | 654 | 58.8% | 399 | 47.0% | 640 | 56.0% | 579 | 52.8% |
| Yes | 458 | 41.2% | 449 | 53.0% | 503 | 44.0% | 518 | 47.2% |
| Impact on standard of living |  |  |  |  |  |  |  |  |
| Worsened | 332 | 29.9% | 317 | 37.4% | 354 | 31.0% | 352 | 32.1% |
| Same or improved | 780 | 70.1% | 531 | 62.6% | 789 | 69.0% | 757 | 69.0% |
| Total | 1112 |  | 848 |  | 1143 |  | 1098 |  |

Note: weighted frequencies.

Lockdown features and levels were combined into pairwise choice tasks using a D-efficient design.[23,24] The design results in 24 tasks. Respondents were allocated randomly to one of the three survey versions, each with eight tasks. Respondents were asked to choose between two lockdown descriptions (Figure 2). The order of the eight tasks was randomised for each respondent to minimise ordering effects.[25]

**Figure 2:**
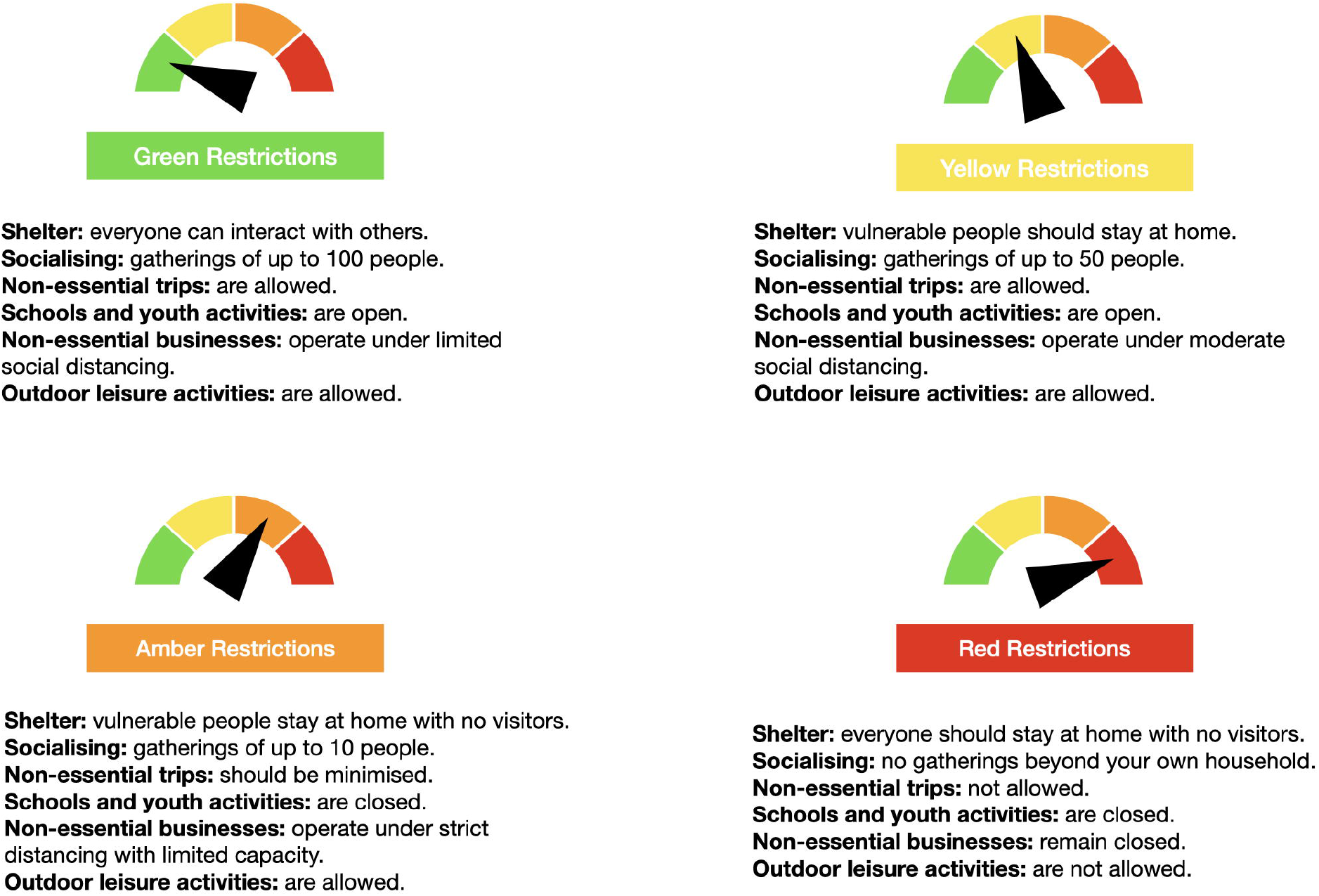

### Patient and Public Involvement

Adult members of the public were invited, using two targeted social media campaigns, to take part in the study development stage (see online Supplemental Figures 1–5). These engagements were used to create the survey’s content and format, and to construct the framing of the Discrete Choice Experiment’s features and levels (see online Supplemental Material document). Twenty-three think-aloud interviews were carried out between the months of June and August 2020. The outcome of each interviews was used iteratively, until saturation was achieved, to make edits to the survey to ensure it captured the intended preferences, was understandable, and minimised respondent burden. The study results will be disseminated to the wider public, with the help of the SAG, using layperson summaries and multimedia content through mass media. Furthermore, the study’s Stakeholder Advisory Group (SAG), which includes a member of Scotland’s Chief Scientist Office’s Public Engagement Group, has been involved since its conception and provided insight into the research questions, overall design and dissemination strategy. Because of the study’s ethical approvals, it is not possible for us to contact the members of the public who took part in the survey development stage, nor respondents of the main survey, to disseminate results individually.

### Statistical Analysis

The devolved governments of the UK set their own lockdown policies; therefore, statistical analysis was conducted separately for each of the four devolved nations of the UK. The minimal sample size for the DCE given the eight tasks per respondent, a baseline choice probability of 50% (given there were two options in each choice set), an accuracy level of 90% and a confidence level of 95%, using Louviere’s formula for choice proportions, was 49 respondents.[22] Given that we aimed to estimate preferences using flexible logit models, we aimed for a conservative size of 1000 per nation in the UK.

We first test if any respondents were unwilling to accept an increase in excess deaths for improvements in other features. This was defined as respondents who always chose the description with the lowest number of excess deaths. The response pattern for these respondents is shown in the online Supplemental Table 2. We estimated a logit regression model to understand the characteristics of this group for each nation. Predictors included: sex, age, self-perceived health, number of children in household, household income quintile, whether they were asked to shield during previous lockdowns, had their main job impacted (furloughed, reduced hours or made redundant), had caring responsibilities and if they had seen their standard of living worsened during the COVID-19 pandemic.

**Table 2.** Characteristics associated with respondents that always minimises excess deaths in the discrete choice experiment tasks

|  |  | England |  |  | Northern Ireland |  |  | Scotland |  |  | Wales |  |  |
| --- | --- | --- | --- | --- | --- | --- | --- | --- | --- | --- | --- | --- | --- |
|  |  | OR | (95% CI) | p value | OR | (95% CI) | p value | OR | (95% CI) | p value | OR | (95% CI) | p value |
| Sex | Female | 1 | (ref) |  | 1 | (ref) |  | 1 | (ref) |  | 1 | (ref) |  |
|  | Male | 0.82 | (0.58 – 1.16) | 0.27 | 0.81 | (0.55 – 1.18) | 0.27 | 1.04 | (0.75 – 1.44) | 0.83 | 1.09 | (0.78 – 1.51) | 0.62 |
| Age | 18-34 | 1 | (ref) |  | 1 | (ref) |  | 1 | (ref) |  | 1 | (ref) |  |
|  | 35-55 | 1.60 | (1.00 – 2.53) | 0.05 | 1.54 | (0.93 – 2.56) | 0.09 | 0.93 | (0.61 – 1.41) | 0.73 | 0.78 | (0.52 – 1.19) | 0.25 |
|  | 55+ | 1.27 | (0.80 – 2.02) | 0.32 | 1.29 | (0.75 – 2.23) | 0.36 | 1.33 | (0.86 – 2.06) | 0.20 | **0.63** | **(0.40 – 0.98)** | **0.04** |
| Health | Very good | 1 | (ref) |  | 1 | (ref) |  | 1 | (ref) |  | 1 | (ref) |  |
|  | Good | 1.26 | (0.77 – 2.04) | 0.36 | 1.07 | (0.62 – 1.84) | 0.81 | 1.01 | (0.64 – 1.58) | 0.97 | 1.31 | (0.84 – 2.03) | 0.23 |
|  | Fair | 1.52 | (0.90 – 2.59) | 0.12 | 0.92 | (0.50 – 1.71) | 0.80 | **1.82** | **(1.11 – 2.97)** | **0.02** | 0.85 | (0.51 – 1.42) | 0.53 |
|  | Bad | 1.36 | (0.59 – 3.10) | 0.47 | 1.06 | (0.46 – 2.46) | 0.89 | 1.20 | (0.53 – 2.72) | 0.66 | 0.79 | (0.35 – 1.81) | 0.58 |
|  | Very bad | 2.16 | (0.67 – 6.95) | 0.20 | 0.82 | (0.19 – 3.63) | 0.80 | 0.28 | (0.02 – 3.98) | 0.35 | 1.56 | (0.32 – 7.62) | 0.58 |
| Shield | No | 1 | (ref) |  | 1 | (ref) |  | 1 | (ref) |  | 1 | (ref) |  |
|  | Yes | 1.18 | (0.80 – 1.74) | 0.41 | 1.07 | (0.70 – 1.64) | 0.76 | 0.97 | (0.63 – 1.51) | 0.91 | 1.25 | (0.85 – 1.85) | 0.26 |
| Adults in household | 1 | 1 | (ref) |  | 1 | (ref) |  | 1 | (ref) |  | 1 | (ref) |  |
|  | 2 | 1.09 | (0.71 – 1.67) | 0.69 | 1.15 | (0.70 – 1.88) | 0.58 | 1.06 | (0.72 – 1.57) | 0.77 | 0.75 | (0.50 – 1.11) | 0.15 |
|  | 3 | 1.19 | (0.66 – 2.16) | 0.56 | 1.14 | (0.61 – 2.13) | 0.69 | 1.03 | (0.58 – 1.82) | 0.93 | 0.57 | (0.31 – 1.06) | 0.08 |
|  | >3 | 1.13 | (0.56 – 2.28) | 0.74 | 1.65 | (0.77 – 3.55) | 0.20 | 1.43 | (0.66 – 3.08) | 0.37 | 1.07 | (0.52 – 2.20) | 0.86 |
| Children in household | 0 | 1 | (ref) |  | 1 | (ref) |  | 1 | (ref) |  | 1 | (ref) |  |
|  | 1 | 0.90 | (0.54 – 1.48) | 0.67 | 0.65 | (0.35 – 1.21) | 0.17 | 0.90 | (0.55 – 1.47) | 0.68 | 0.97 | (0.60 – 1.58) | 0.91 |
|  | >2 | 1.61 | (0.68 – 3.83) | 0.28 | 2.21 | (0.94 – 5.21) | 0.07 | 0.43 | (0.09 – 1.97) | 0.28 | 0.65 | (0.22 – 1.89) | 0.42 |
| Household income |  |  |  |  |  |  |  |  |  |  |  |  |  |
|  | £0 - £10,400 | 1 | (ref) |  | 1 | (ref) |  | 1 | (ref) |  | 1 | (ref) |  |
|  | £10,400 - £20,800 | 1.48 | (0.76 – 2.92) | 0.25 | 1.51 | (0.74 – 3.11) | 0.26 | 0.69 | (0.39 – 1.22) | 0.20 | **0.49** | **(0.29 – 0.83)** | **0.01** |
|  | £20,800 - £31,200 | 1.48 | (0.74 – 2.98) | 0.27 | 1.76 | (0.87 – 3.58) | 0.12 | **0.54** | **(0.31 – 0.95)** | **0.03** | **0.57** | **(0.34 – 0.96)** | **0.04** |
|  | £31,200 - £52,000 | 1.30 | (0.65 – 2.60) | 0.46 | 2.01 | (0.98 – 4.11) | 0.06 | 0.68 | (0.39 – 1.19) | 0.17 | 0.86 | (0.26 – 0.94) | 0.57 |
|  | £52,000+ | 1.38 | (0.65 – 2.93) | 0.40 | 1.48 | (0.63 – 3.47) | 0.36 | 0.88 | (0.49 – 1.61) | 0.69 | **0.49** | **(0.79 – 1.56)** | **0.03** |
| Education |  |  |  |  |  |  |  |  |  |  |  |  |  |
|  | Less than higher education | 1 | (ref) |  | 1 | (ref) |  | 1 | (ref) |  | 1 | (ref) |  |
|  | Higher education degree | 1.29 | (0.92 – 1.83) | 0.142 | 0.95 | (0.64 – 1.42) | 0.81 | **1.77** | **(1.28 – 2.45)** | **<0.01** | 1.11 | (0.79 – 1.56) | 0.53 |
| Job impact |  |  |  |  |  |  |  |  |  |  |  |  |  |
|  | No | 1 | (ref) |  | 1 | (ref) |  | 1 | (ref) |  | 1 | (ref) |  |
|  | Yes | 0.80 | (0.53 – 1.22) | 0.307 | **0.58** | **(0.35 – 0.97)** | **0.04** | **0.62** | **(0.40 – 0.95)** | **0.03** | 0.93 | (0.63 – 1.37) | 0.73 |
| Caring responsibility |  |  |  |  |  |  |  |  |  |  |  |  |  |
|  | No | 1 | (ref) |  | 1 | (ref) |  | 1 | (ref) |  | 1 | (ref) |  |
|  | Yes | 0.81 | (0.48 – 1.36) | 0.424 | 1.23 | (0.76 – 1.97) | 0.40 | 0.71 | (0.45 – 1.13) | 0.15 | 1.33 | (0.88 – 1.99) | 0.17 |
| Affected usual healthcare |  |  |  |  |  |  |  |  |  |  |  |  |  |
|  | No | 1 | (ref) |  | 1 | (ref) |  | 1 | (ref) |  | 1 | (ref) |  |
|  | Yes | 1.00 | (0.71 – 1.42) | 0.987 | 1.07 | (0.73 – 1.57) | 0.73 | 1.05 | (0.76 – 1.45) | 0.76 | 0.84 | (0.60 – 1.17) | 0.29 |
| Impact on standard of living |  |  |  |  |  |  |  |  |  |  |  |  |  |
|  | Same or improved | 1 | (ref) |  | 1 | (ref) |  | 1 | (ref) |  | 1 | (ref) |  |
|  | Worsened | 0.99 | (0.66 – 1.47) | 0.949 | 1.13 | (0.74 – 1.73) | 0.56 | 1.16 | (0.81 – 1.67) | 0.41 | 0.95 | (0.66 – 1.37) | 0.78 |

Note: **bold** indicates significance at the 95% level.

We then analysed the choice tasks using an errors-component logit model, allowing for the panel structure of the data.[26] Parameter estimates represented the effect of each feature on preferences. The ratio of estimates represents the trade-off between two features. Further, trade-offs between different features, when elicited in terms of a common denominator, can be added to estimate the overall trade-off for a particular lockdown scenario. When elicited in terms of excess deaths, these trade-offs indicate the maximum number of lives that need to be saved to introduce a hypothetical lockdown scenario. For example, how many excess deaths would need to be saved when introducing a four-week strict lockdown that cancels all non-COVID-19 healthcare procedures?

The difference in trade-offs between two lockdown scenarios can be interpreted as the maximum number of excess deaths that would be accepted if the more preferred scenario were introduced. To illustrate how these differences can inform policy, we assume that each nation faces a four-week red level (see Figure 1) restriction lockdown that postpones all non-COVID-19 healthcare procedures, and estimate the acceptable number of excess deaths to have this eased to less strict lockdown scenarios. Specifically we compare easing to 12 different lockdowns made up of combinations of amber and yellow restrictions (Figure 1) that vary in length between 8,10 and 12 weeks, and in whether they postpone healthcare services.

Data was weighted to ensure a representative sample in terms of age and sex using iterative proportional fitting.[27] All logit models were estimated using maximum likelihood techniques using the statistical software R (version 3.6.3). Standard errors and confidence intervals (CIs) were computed using the delta method.

## Results

4120 respondents completed the survey: 1112 in England, 848 in Northern Ireland, 1143 in Scotland, and 1098 in Wales. Table 1 shows the sample descriptive characteristics across nations.

The number of respondents who consistently chose the alternative with the least excess deaths was 225 (20.2%) in England, 193 (22.8%) in Northern Ireland, 262 (22.9%) in Scotland, and 247 (22.5%) in Wales. Results from the logit model are shown in Table 2. In England, none of the considered variables were associated with respondents always choosing the lowest number of excess deaths. In Northern Ireland, this response pattern was negatively associated with respondents who experienced an impact on employment (adjusted odds ratio [OR] 0.58 [95% CI 0.35–0.97], p=0.04). In Scotland, this response pattern was also negatively associated with respondents who experienced an impact on employment (0.62 [0.40–0.95], p=0.03), and household income of £20,800-£31,200 compared to the reference level of £0-£10,400 (0.54 [0.31–0.95], p=0.03). Furthermore, this response pattern was positively associated with having a higher education degree (compared to less than higher education) (1.77 [1.28–2.45], p<0.01) and fair self-reported health compared to very good (1.82 [1.11–2.97], p=0.02). In Wales, this response pattern was negatively associated with age over 55 compared to 18-34 (0.63 [0.40–0.98], p=0.04), household incomes of £10,400-£20,800 (0.49 [0.29–0.83], p=0.01), £20,800-£31,200 (0.57 [0.34–0.96], p=0.04) and over £52,000 (0.49 [0.79–1.56], p=0.03) compared to £0-£10,400. Univariate analyses for each factor are shown in the online Supplemental Table 3.

**Table 3.**
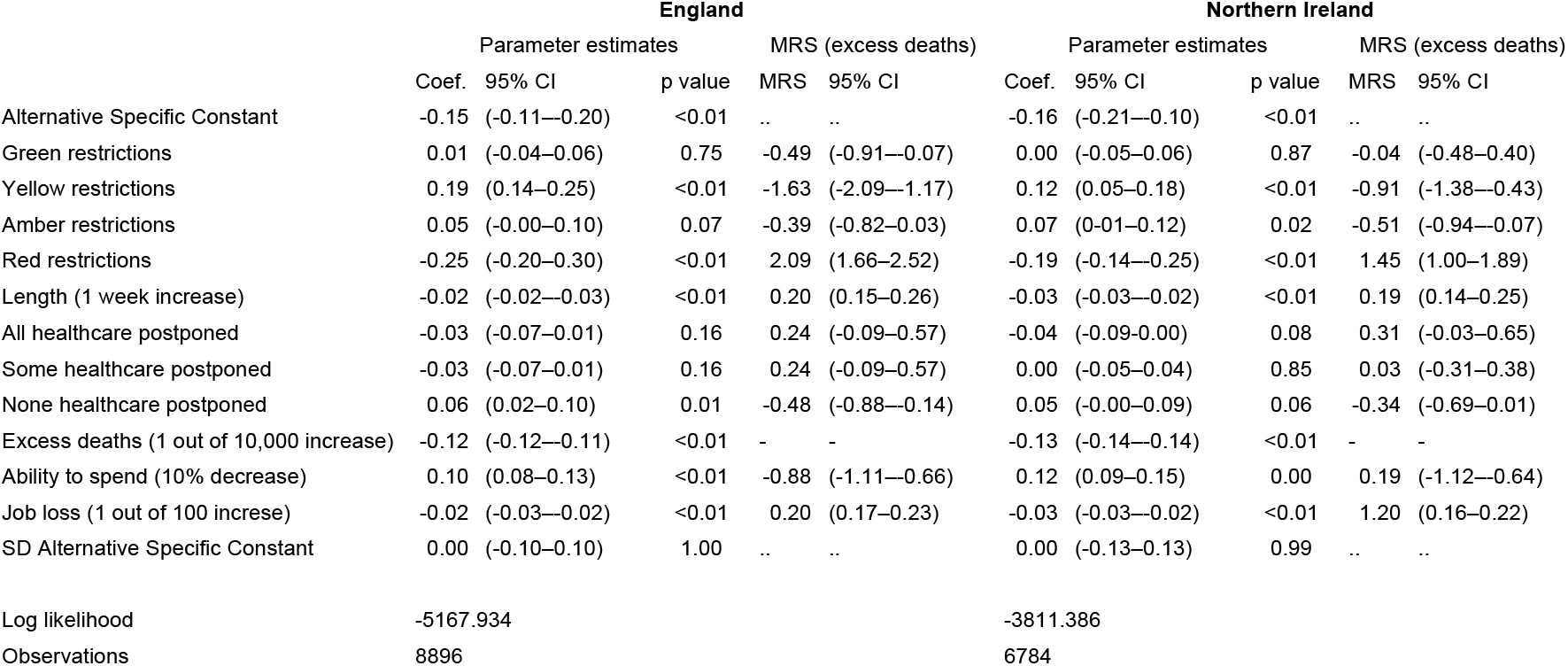
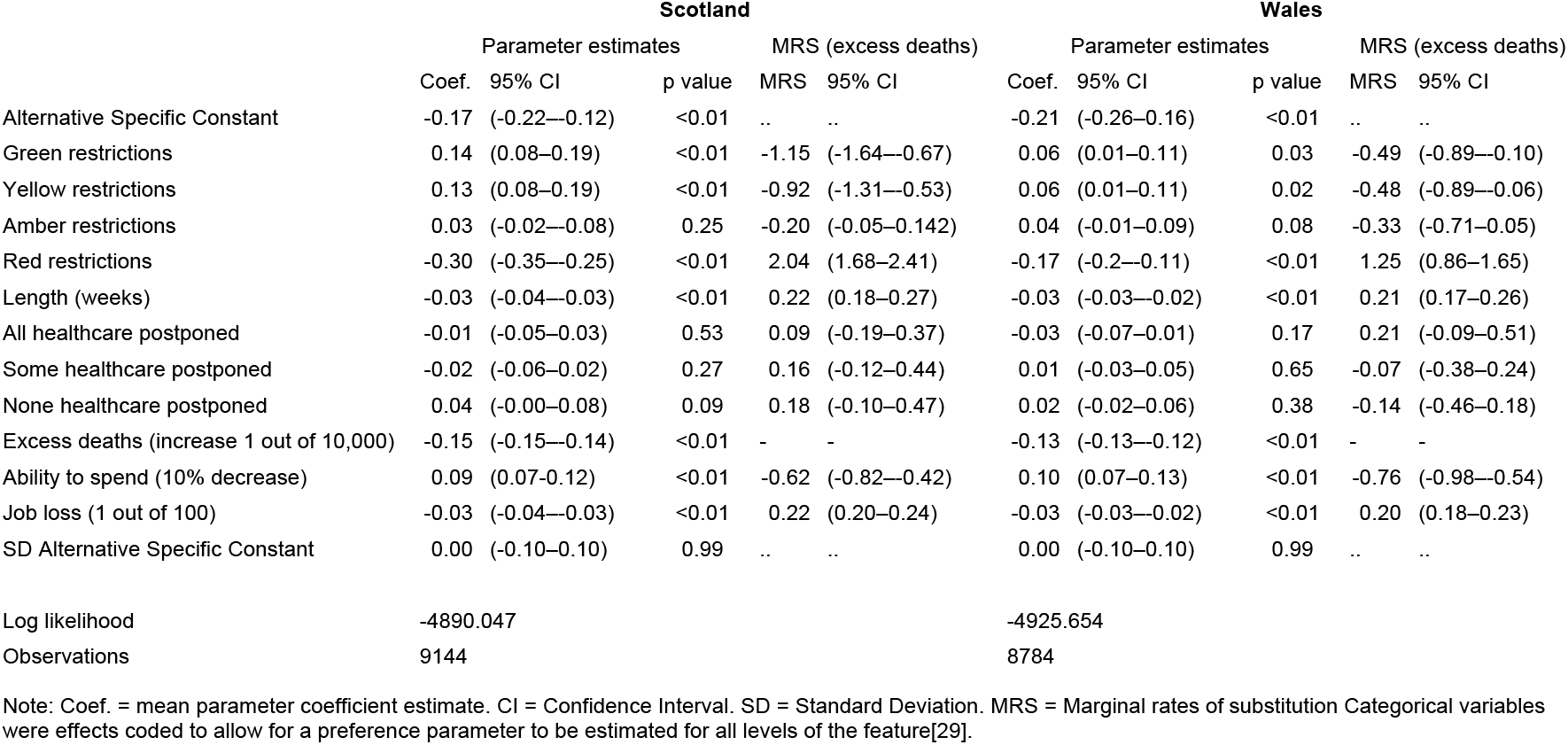
Preferences for lockdown features.

The preference parameter estimates and corresponding trade-offs in terms of excess deaths based on responses to the choice tasks are shown in Table 3. Across the four nations, respondents prefer lockdowns with less strict restrictions (i.e., green and yellow level) to strict ones (i.e., amber and red level), shorter lockdowns, fewer excess deaths, fewer job losses, and less impact on their ability to buy goods. In England, Northern Ireland and Scotland, respondents prefer no postponement of routine healthcare procedures (at the 10% level). The maximum number of lives (out of 10,000) that need to be saved to accept a change in each of the lockdown features and consequences is shown in the MRS column for each nation.

Figure 3 shows the acceptable maximum excess deaths for easing restrictions from a further 4-week red lockdown to the less strict lockdowns. The highest aversion to strict lockdowns is found in England, followed by Scotland, Northern Ireland and Wales, as seen by the higher number of acceptable excess deaths for lockdown easing. For example, the maximum number of acceptable deaths when easing to an 8-week yellow restriction with no healthcare postponement is 3.62 (95% CI 2.67–4.58) in England, 2.22 (1.21–3.24) in Northern Ireland, 2.41 (1.57–3.24) in Scotland, and 1.10 (0.18–2.02) in Wales. These rates equal 18958, 361, 1265, and 323 excess deaths for each nation, respectively.

**Figure 3:**
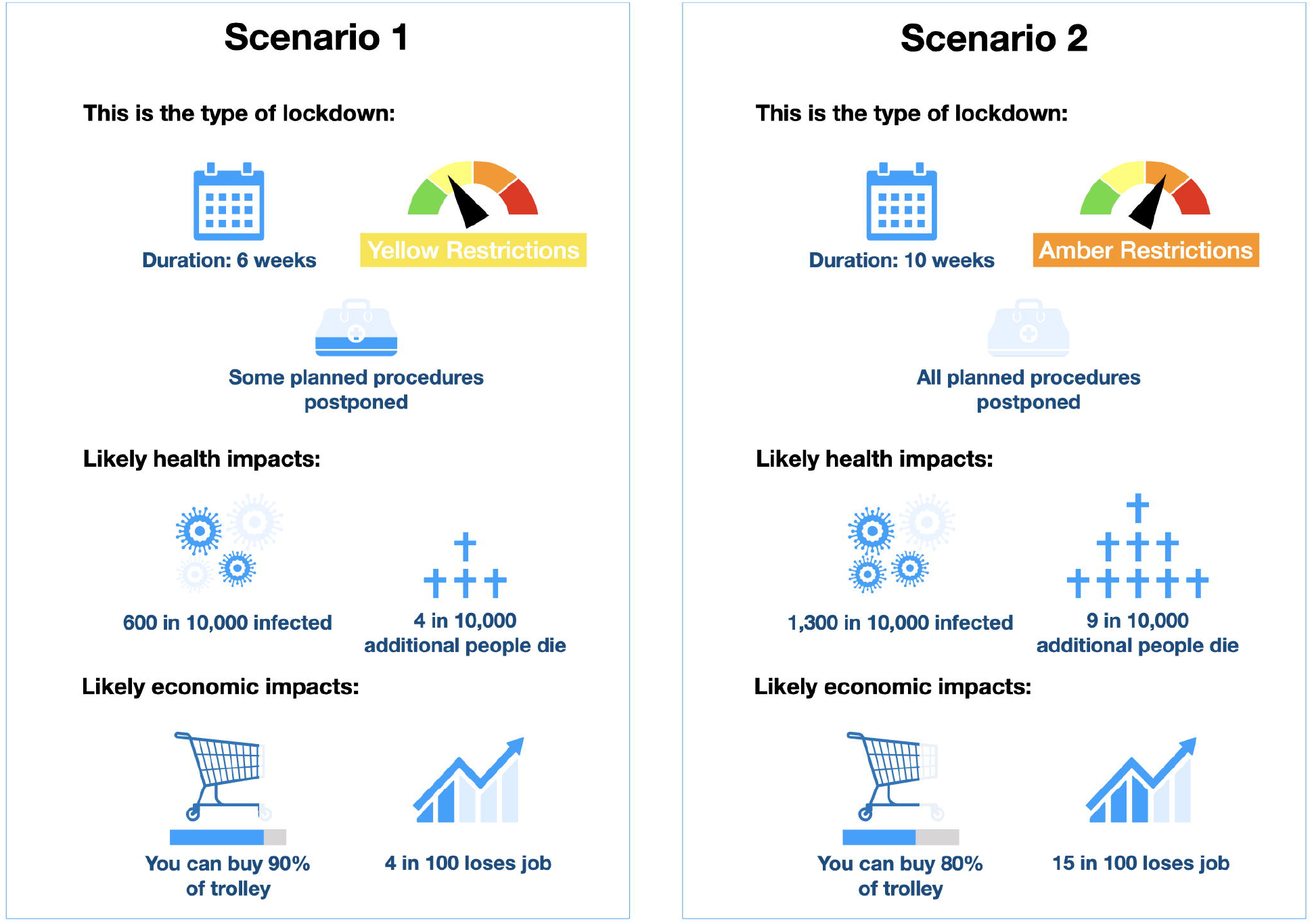

As expected, the maximum number of acceptable deaths is lower when moving to more strict (e.g., amber over yellow) and longer lockdowns that postpone routine healthcare procedures. For example, the difference in the acceptable number of deaths between a 4-week red lockdown and a 12-week amber lockdown with healthcare postponement is 0.85 (0.03–1.67) in England and not statistically different from zero in Northern Ireland (*X*^*2*^*=0*.*88*, p=0.35), Scotland (*X*^*2*^*=1*.*84*, p=0.17), and Wales (*X*^*2*^*=0*.*08*, p=0.77). This suggests that respondents in Northern Ireland, Scotland, and Wales are indifferent between continuing with a further 4-week red restriction and easing to a 12-week amber restriction with healthcare postponement.

## Discussion

The elicitation of public values and trade-offs for different lockdown features can help guide government policies during a pandemic. We found evidence that four out of five respondents were willing to accept an increase in excess deaths for relaxations in lockdown restrictions. This suggests that as the governments of the devolved nations consider easing lockdown, the public will be willing to accept an increase in excess deaths. We also estimated acceptable excess deaths for such relaxations.

With the roll-out of pharmaceutical interventions and the increase in data available to model the impact of changes in restrictions, our model can help inform policy makers about what lockdown policies are acceptable given the estimated trade-offs. We found that respondents in England are the most averse to the introduction of short *circuit-breaker*-type lockdowns, thus accepting a higher number of excess deaths to avoid them. In contrast, these strict lockdowns were more palatable to respondents in Wales.

Trade-off values can also be interpreted as the number of lives that need to be saved if a less preferred and expectedly stricter lockdown is implemented. Our model can be used to assess whether the expected health benefits in terms of a reduction in the number of excess deaths outweigh costs in terms of increased restrictions. As an example, modelling by Ferguson et al. (2020) contended that a one-week earlier strict lockdown in England during COVID-19’s first wave would have saved 20,000 lives.[28] Our findings suggest that the number of acceptable deaths in England for a one-week strict (red level restrictions) lockdown is 2.53 out of 10,000, or 14,170 lives, which is less than the number of lives that would have been saved (see online Supplemental Material p.14 for details). Thus, based on these results, the benefits of introducing an earlier lockdown would have outweighed the costs in terms of lockdown restrictions. These insights can be useful as UK governments consider easing lockdown restrictions or the introduction of new ones if future infection waves occur.

Whilst we limited our analysis to consider acceptable excess deaths, a strength of our model is that it can be used to determine value in terms of other features included, i.e. acceptable reductions in spending or job losses associated with a particular lockdown scenario. We found that respondents in Scotland were less sensitive to losses in their own spending ability compared to other nations. For example, the average acceptable loss in spending ability for a four-week red level lockdown in Scotland is 49%, while in England it is 36%, Northern Ireland it is 29%, and Wales 30%. A detailed calculation of these MRS can be found in the online Supplemental Material (p. 16). Thus, a targeted compensation instrument could target other economic consequences, such as joblessness, in Scotland and consumer spending ability in the other nations.

Our study is not exempt from limitations. A potential limitation is that individual’s preferences regarding the features of lockdown may be evolving. Until March 2020, respondents would not have experienced a lockdown. However, we conducted our survey in October-December 2020, hence all respondents would have experience of the first lockdown. The study was, however, conducted before the second lockdown.

The dynamics of preferences and trade-offs for lockdown should be closely monitored. Another possible limitation is that we identified respondents as excess death minimisers if they chose the option with the minimum number of deaths in all eight choice tasks. This response pattern could also represent a decision-heuristic for respondents to complete the tasks. It is also possible that respondents are considering excess deaths, and trading, but the combination of feature levels results in the option with the lowest number of excess deaths. Either way, this suggests our estimate of 80% of respondents being willing to trade would be an underestimate. Further, we estimated trade-offs across the entire sample, allowing for the possibility that such responders were traders. We have not attempted to explain preference heterogeneity across nor within nations. Our study did not look at the relative importance of the different dimensions of lockdown restrictions (shelter, socialising, non-essential trips, schools and youth activities, non-essential businesses and outdoor activities). Future work could use a DCE to explore this; given current discussions around international travel, this dimension could be included. We focused on the preferences of the public; future research could explore the preferences of policy makers and health professionals.

## Conclusions

In this study we have provided new insight into preferences for lockdown policies across the four UK nations using a DCE. The majority of respondents from all four devolved nations were willing to accept an increase in excess deaths for relaxation in lockdown restrictions. Respondents from England were more willing to accept an increase in excess deaths, followed by Scotland, Northern Ireland and Wales. Our model can also be used to estimate the reduction in excess deaths required to justify increasing lockdown restrictions. Whilst we focused on excess deaths, trade-offs could also be estimated in terms of acceptable changes in spending power and job losses, as well as combinations of these features. Such analysis will help identify which levers best support lockdown strategies whilst maintaining public confidence and maximising compliance.

## Data Availability

Anonymised cross-sectional data from the analysis can be made available by the corresponding author after the authors' review of reasonable requests. The published protocol can be found at: https://bmjopen.bmj.com/content/10/11.

## Acknowledgements

We want to thank the members of the public who took part in the think aloud interviews as part of the study development stage. We thank the members of the Stakeholder Advisory Group for their continued involvement in the research study. We also thank Dr Dwayne Boyers for his internal review of the study protocol and Professor Vikki Entwistle for comments on the protocol.

## Contributors

MR, VW, MG, RAS & LEL-R conceptualised the study, contributed to the overall design of the survey experiment and contributed to the interpretation of the data. MG and RAS carried out the think-aloud interviews as part of the developmental work. LEL-R undertook the analysis, including the R programming of the statistical models and is the study’s guarantor. MR reviewed the statistical model and contributed to the analysis of the data. SP and DP contributed comments to the development of the protocol, and discussion of public health implications and helped shape the overall interpretation. All authors approved the final protocol. All authors had access to all the data, contributed to the writing of the paper and had final responsibility for the decision to submit for publication. The corresponding author attests that all listed authors meet authorship criteria and that no others meeting the criteria have been omitted.

## Ethical approval

The study received ethical approval from the University of Aberdeen’s College of Life Sciences and Medicine Ethical Review Board (Reference CERB/2020/6/1974). All participants provided informed consent.

## Dissemination to participants and related patient and public communities

The results have been and will be presented at national and international conferences. Dissemination plans to inform the community of this study’s results include social media and University’s newsletter. Authors will liaison with the study’s Stakeholder Advisory Group to ensure maximum policy impact of the study’s findings.

## Transparency

The corresponding author affirms that this manuscript is an honest, accurate, and transparent account of the study being reported; that no important aspects of the study have been omitted; and that any discrepancies from the study as planned (and, if relevant, registered) have been explained.

