## Supplemental Material for "Public acceptability of non-pharmaceutical interventions to control a pandemic in the United Kingdom: a discrete choice experiment"

### Online Supplementary Material

**Supplemental Table 1.** Features and levels used in the discrete choice experiment.

| Feature | Description | Levels |
|---|---|---|
| Type of Lockdown (Severity of restrictions) | How restrictive the lockdown (based on a colour/tier system). | Green<br>Yellow<br>Amber<br>Red |
| Length | How long the lockdown is in place. | 3 weeks<br>6 weeks<br>10 weeks<br>16 weeks |
| Postponement of usual non-medical care | Whether non-pandemic medical care is postponed. | No procedures are postponed<br>Some procedures are postponed<br>All procedures are postponed |
| Excess deaths | Number of excess deaths (expressed as a fraction of 10,000). | 1<br>4<br>9<br>13 |
| Infections [a] | Number of infections (expressed as a fraction of 10,000). | 100<br>600<br>1,300<br>2,000 |
| Ability to buy things | How much of the goods that respondents are able to buy today will they be able to buy in a year's time. | 100% of their shopping trolley<br>90% of their shopping trolley<br>80% of their shopping trolley<br>70% of their shopping trolley |
| Job losses | How many people lose their job (expressed as a fraction of 100). | 0<br>4<br>15<br>25 |

Note: [a] Number of infections were linked to the excess death feature using an Infection Fatality Rate of 0.7%.

### Discrete choice experiment: think-aloud developmental work.

Virtual think-aloud (TA) interviews were conducted using MS Teams with colleagues from the University of Aberdeen (n=10) and members of our Stakeholder Advisory Group (n=4). Subsequently, we recruited members of the general public to participate in virtual TAs via two Facebook recruitment campaigns. Facebook users, resident in the United Kingdom and over 18 years of age, were shown an advertisement inviting them to participate in a study about COVID-19 lockdowns. Supplemental Figure 1 shows the advertisement for the first campaign.

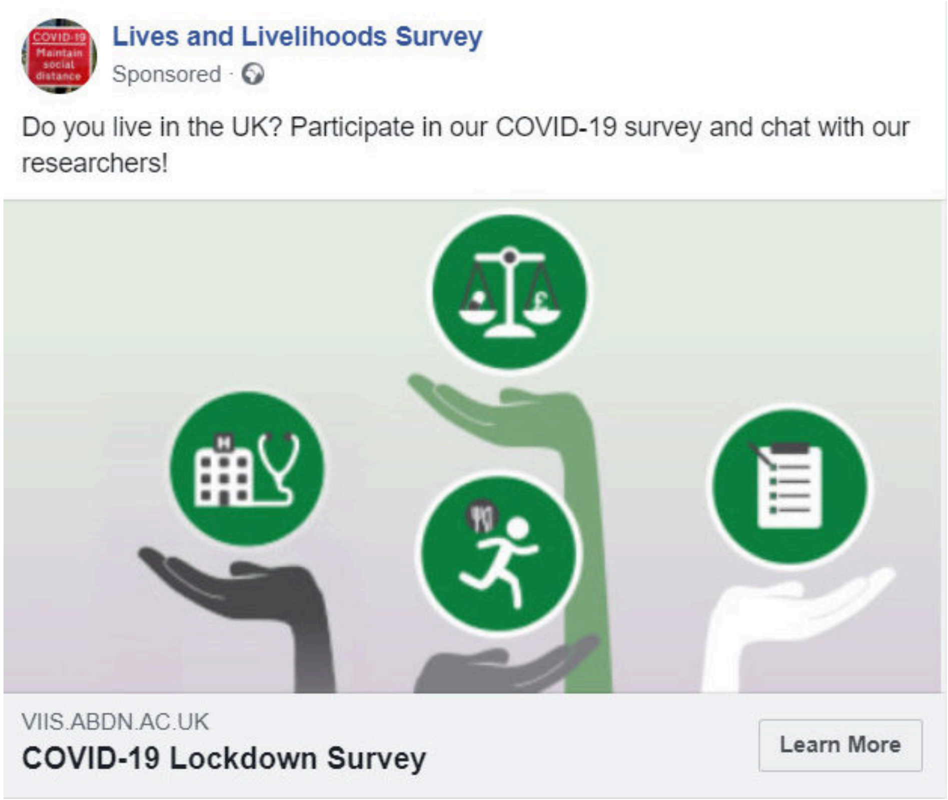


**Supplemental Figure 1.** Facebook campaign 1, advertisement appearance.

Upon clicking the advertisement, users were directed to a landing page with more information and were asked to enter their names and email addresses in a web form to indicate their interest in participating in an interview. Supplemental Figure 2 shows the landing page.

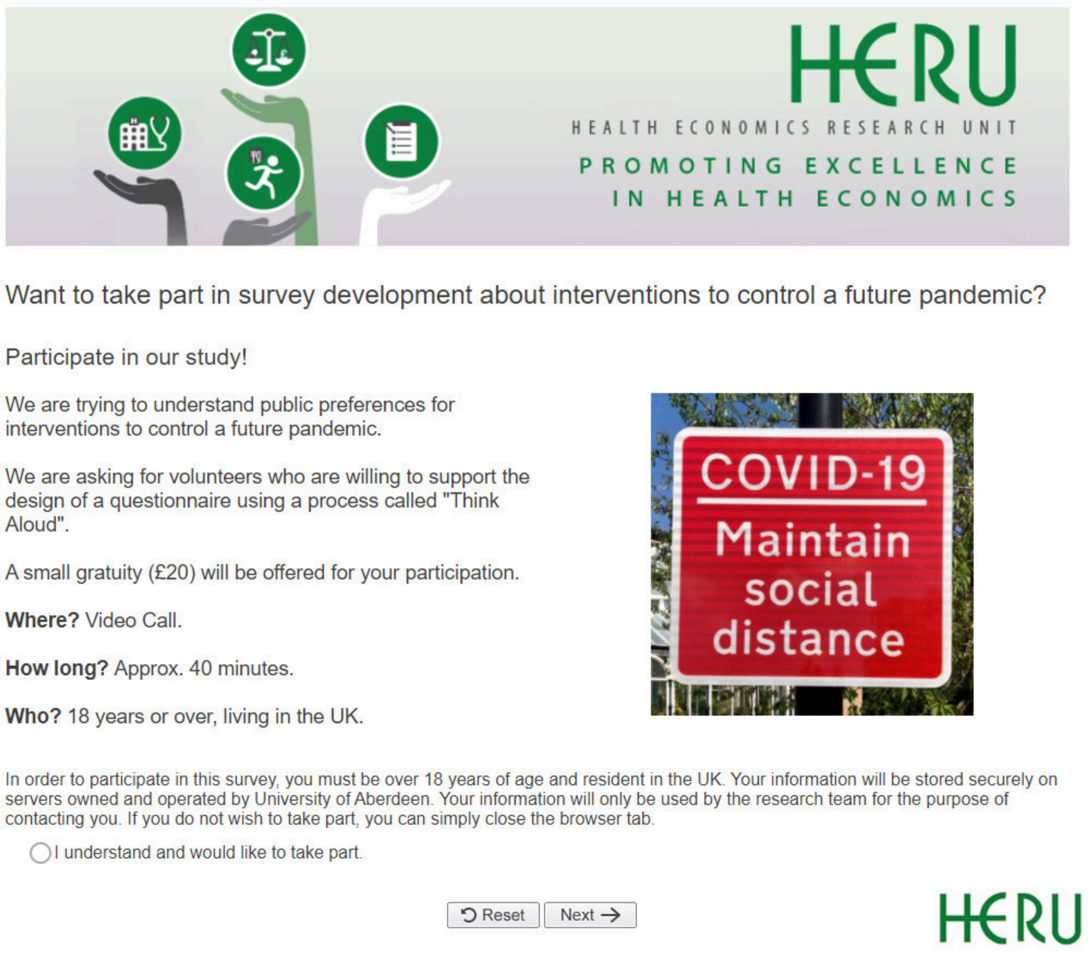


**Supplemental Figure 2.** Participant Landing page

The first campaign ran from August 8, 2020, until August 14, 2020, was shown to 11,632 users and resulted in 343 clicks on the advert. Whilst 32 respondents indicated interest in participating by submitting their contact information through the landing page, only a limited number responded to contact by the researcher. To improve uptake we modified the Facebook advertisement, including information on the £20 voucher participants would receive for their participation (Supplemental Figure 3). The campaign with the modified text ran from August 25, 2020, until August 31, 2020, was shown to 10,912 and resulted in 291 clicks. 52 respondents indicated an interest for an interview by submitting their contact information through the landing page. Again, not all respondents who indicated an interest in participation via the landing page responded to the researcher’s contact. In total 23 interviews were conducted from across the two campaigns.

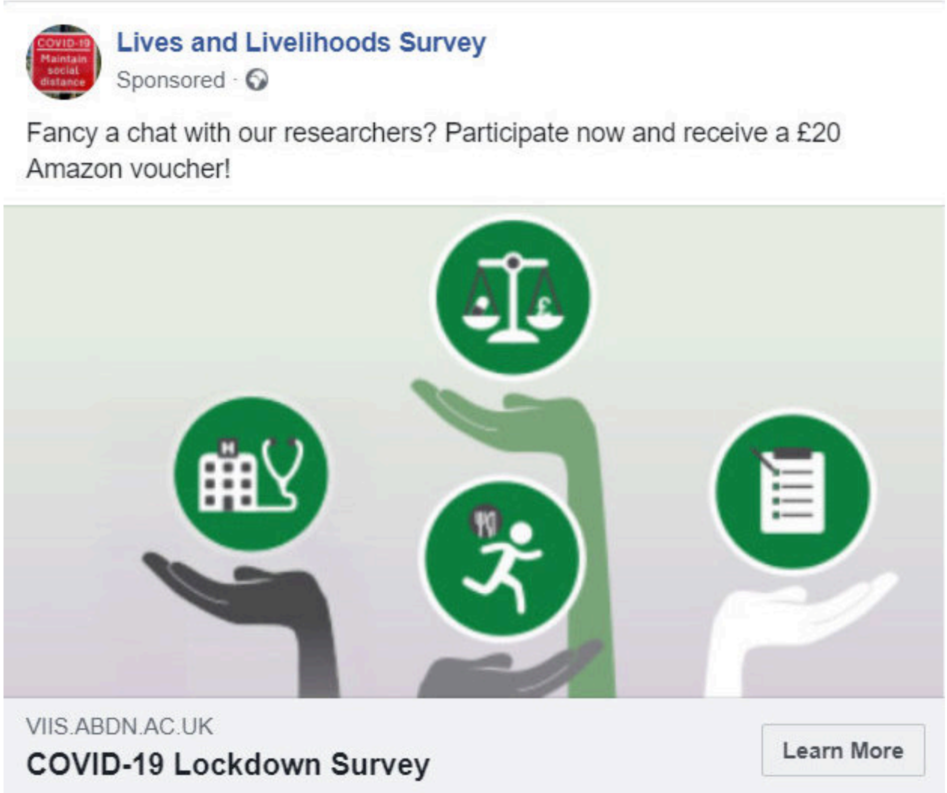


**Supplemental Figure 3.** Facebook campaign 2, advertisement appearance.

Facebook does not offer control over the demographic composition of users targeted by the ad beyond general inclusion and exclusion criteria. We specified our target group as users resident in the UK over 18 years of age. The demographics of Facebook users that were shown the advertisement skewed older and female. Supplemental Figure 4 shows the demographics for campaign 1, and Supplemental Figure 5 shows the demographics for campaign 2.

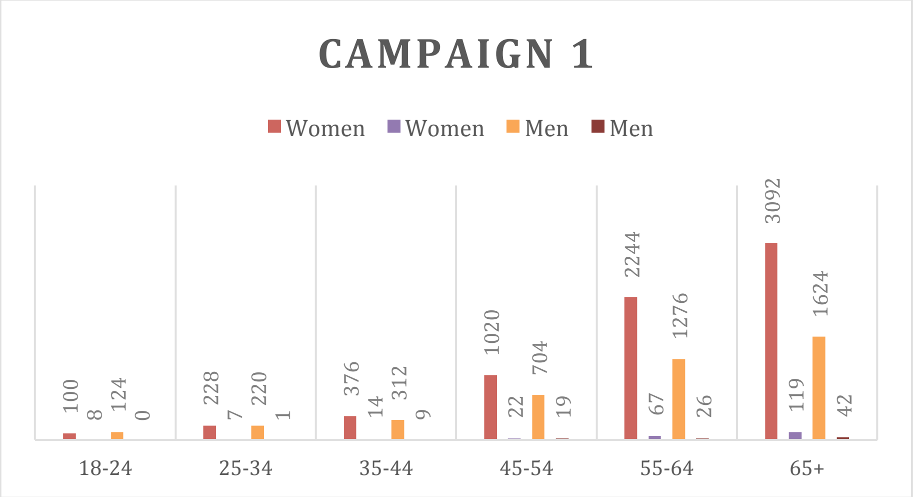


**Supplemental Figure 4.** Campaign 1 demographics

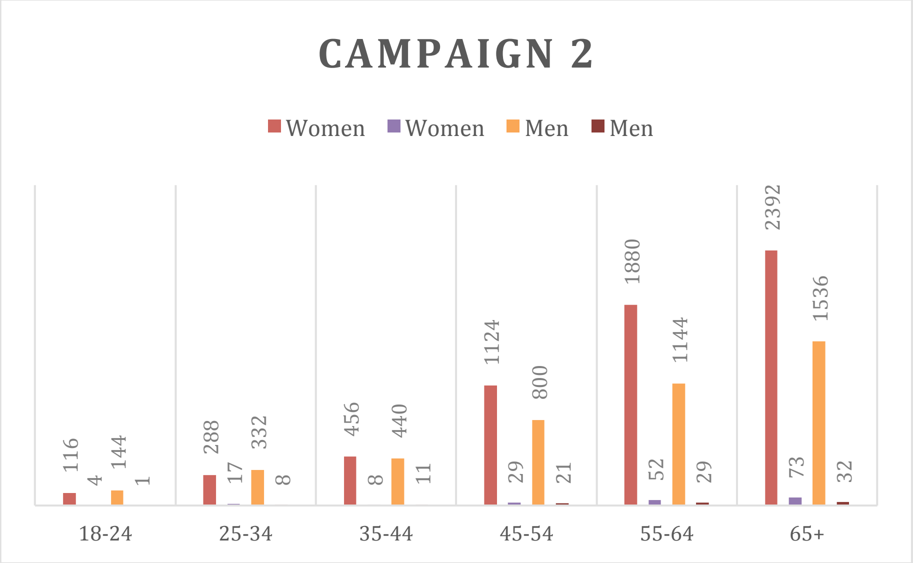


**Supplemental Figure 5.** Campaign 2 demographics

For all TAs, participants were asked to share their device's screen with the interviewers and verbalise their thought processes whilst responding to the survey. As a warm-up, they were asked to think aloud whilst responding to the question: "*How many windows are there in your house?"* Respondents were told to consider the interviewer as a silent observer of their thought process. Interviewers did, however, encourage respondents to verbalise their thoughts if they were silent for a short period. Respondents were told there were no right or wrong answers. The interviews lasted between 45 and 90 minutes.

A number of changes were made to the DCE survey as a result of participant feedback.

1. Presentation of the excess death, number of infections, and job loss features

In the TA interview used for internal testing, the features for excess death, number of infections, and job losses were presented differently. The number of jobs lost and the number of people infected were presented as fractions of 100. In contrast, the excess death feature was presented as absolute numbers of additional people dying over the expected figure during a normal year. This led to the excess death feature dominating the choices of a considerable number of participants, with some participants stating that they ignored all other features and only considered the number of excess deaths presented in the choice task.

While this might be an expression of a valid preference, the feedback we received included evidence that the presentation of the excess death feature in absolute numbers inflated its importance relative to other features. One participant stated that, while they recognised that job loss was presented as fractions, in their mind, they ignored the denominator of the job loss feature and directly compared its numerator to the absolute figures presented for the excess death feature.

We thus changed the presentation of excess deaths and number of infections to be uniform across the choice task. In the final survey, the number of infections and excess deaths are presented as fractions of 10,000.

2. Presentation and placement of lockdown restrictions feature

In the TA interview for internal testing, the colour-coded visual for the lockdown restrictions was prominently presented at the top of each choice option. Some participants interpreted the graphic as a summary of the choice option as a whole rather than as an independent feature.

We thus changed the visual position for the lockdown restrictions to appear next to the visual for the lockdown duration feature.

Another contributing factor was that the lockdown restrictions feature was initially presented to participants as “lockdown type”. The group of features representing policy choices (lockdown severity, lockdown length, and postponed procedures) was described in a very similar way as “type of lockdown”.

We renamed the feature to “lockdown restrictions” and changed all visuals to read “(Colour) restrictions” to differentiate more clearly between the “lockdown restrictions” feature and the “lockdown type” group of features.

3. Visual presentation of the number of infections feature

The TA for internal testing displayed a static visual for the number of infections feature that did not change according to the level presented. Several participants stated that a changing visual would improve the presentation of this feature. We thus changed the visual to change with an increasing number of infections.

4. Presentation of the shopping trolley feature

Initially, the text under the visual for the ‘shopping trolley’ feature read “X% of the trolley.” Some participants interpreted this to mean the economic impact on society rather than the economic impact on themselves. We changed the text to read “You can buy X% of the trolley.”

5. Explanation of the shopping trolley feature, warm-up questions for the shopping trolley

Some participants were concerned that the initial explanation of the shopping trolley focused on consumption rather than the general cost of living. One participant expressed concerns that this might not accurately reflect the experiences of impoverished respondents. We expanded the explanation of the shopping trolley feature to include housing costs and utility bills.

The initial warm-up questions presented next to the explanation of the shopping trolley feature referred to respondents' income. Some participants were confused by the question as the explanation for the shopping trolley feature presented the impact in terms of how much respondents could afford to buy. As many respondents reduced consumption during the lockdown, they were unsure how to respond to the question.

We removed references to respondent income from the warm-up questions and instead asked respondents about the impact the pandemic and lockdown measures had on their household's standard of living and how concerned they were about how much their household could afford to buy in a year's time.

6. Warm-up questions for the job loss feature

In an earlier version of the survey, the warm-up question attached to the explanation of the job loss feature asked participants about their concerns about losing their jobs. As this feature was meant to elicit respondents' attitudes from a social-inclusive perspective, we changed the question to read "How concerned are you about rising unemployment as a result of the COVID-19 pandemic?"

7. MFQ20: Likert scale anchors

The initial presentation of the MFQ20 presented the anchors for different points on a 6-point Likert scale ("not at all relevant" to "extremely relevant" and "strongly disagree to "strongly agree") at the top of the page. For the selection matrix, points on the scale were labelled with numbers running from 0-5 to mimic the presentation of the paper-based MFQ 20.

We observed that the top of the page was not visible for participants while answering the questions, leading them to spend much time scrolling up and down on the page. We amended the selection matrix to display the anchors next to the numbered points on the Likert scale.

8. Government performance assessment

Some respondents were confused by the initial wording of the question asking about the performance of the UK government. We changed the question to specify the Westminster government.

9. Thank-you message

One respondent felt that the thank-you message at the end of the survey was not heartfelt enough. We changed the message to acknowledge respondents' efforts and reaffirmed the value of their responses.

10. Ease-of-use updates

To make the survey more engaging, we made various improvements to the interface and presentation formats. This included a progress bar at the top of the screen, mouse-hover explanations for different selection options, and input prompts.

11. Reducing survey completion time

Initially, participants took up to 90 minutes to complete the survey (while verbalising their thoughts). We implemented several improvements to reduce completion time.

We reduced redundant slides reminding participants of the meaning of the feature visuals before starting the DCE. We tested the updated version with TA participants and noticed no adverse effect on participants' ability to understand the task.

An earlier version of the survey featured four warm-up questions attached to the excess death feature. They were presented in two pairs of two 5-point Likert scale questions, asking 1a) how concerned participants were that they could die from COVID-19, 1b) how concerned they were that their loved ones could die from COVID-19, 2a) how concerned they were that they could not access healthcare during the COVID-19 pandemic, and 2b) how concerned they were that their loved ones could not access healthcare during the COVID-19 pandemic. We combined both pairs of questions into two questions asking about participants' concerns about *themselves or loved ones* about 1) the risk of death from COVID-19 and 2) health care access, respectively.

To compensate, we added question asking about the perspective respondents took while completing choice tasks. The question asked whether respondents thought about a) what was best for them, b) what was best for their loved ones, c) what was best for their community, and d) what was best for their country. We conducted a/b testing for two types of questions: one ranking question where respondents indicated the order of importance of the four options, and one question where respondents indicated the most important factor out of the four choices. In accordance with the feedback we received from TA participants, we decided to implement a multiple choice question where respondents could select as many options as needed.

We observed participants struggling with the large number of options for the questions assessing participants' willingness to endure different lockdown restriction levels. Especially on mobile devices such as smartphones and tablets, participants spent much time scrolling through options. We reduced the number of available options in the drop-down menu by removing the odd numbers of weeks.

We reduced the word count of the explanatory messages introducing each new section of the survey. In subsequent TAs, we closely monitored whether this would decrease participants' ability to understand and complete the survey and observed no difference.

### Estimation of lockdown scenario trade-offs using marginal rates of substitution for excess deaths

The calculation for the marginal rate of substitution (MRS) for introducing a 1-week red restriction lockdown where all routine non-COVID healthcare procedures are postponed is the addition of the MRS for the features that describe the scenario, such that:

$$\frac{\beta_{red\_restrictions}}{-\beta_{excess\_deaths}} + \frac{\beta_{length} \times X_{weeks}}{-\beta_{excess\_deaths}} + \frac{\beta_{all\_health\_postponed}}{-\beta_{excess\_deaths}} \quad (1)$$

This can be simplified as:

$$\frac{\beta_{red\_restrictions} + \beta_{length} \times X_{weeks} + \beta_{all\_health\_postponed}}{-\beta_{excess\_deaths}} \quad (2)$$

Following (2), the MRS for the scenario described above are:

England:

$$\frac{-0.246 + (-0.024 \times 1\ ) + (-0.028)}{-0.118} = -2.53$$

Northern Ireland:

$$\frac{-0.192 + (-0.026 \times 1) + (-0.041)}{-0.133} = -1.95$$

Scotland:

$$\frac{-0.298 + (-0.032 \times 1) + (-0.013)}{-0.146} = -2.35$$

Wales:

$$\frac{-0.165 + (-0.028 \times 1) + (-0.028)}{-0.132} = -1.67$$

Standard errors and 95% Confidence Intervals (CI) are calculated using the delta method and are shown below.

| Nation | MRS (absolute) | Standard Error | Lower Confidence Interval | Upper Confidence Interval |
|---|---|---|---|---|
| England | 2.53 | 0.27 | 2.00 | 3.06 |
| Northern Ireland | 1.95 | 0.28 | 1.39 | 2.50 |
| Scotland | 2.35 | 0.23 | 1.89 | 2.81 |
| Wales | 1.67 | 0.26 | 1.17 | 2.18 |

### Estimation of lockdown scenario trade-offs using marginal rates of substitution for decreases in the ability to buy things

The calculation for the marginal rate of substitution (MRS) for introducing a 2-week red restriction lockdown where all routine non-COVID healthcare procedures are postponed in terms of changes (decreases) in the ability to spend are:

England:

$$\frac{-0.246 + (-0.024 \times 4\ ) + (-0.028)}{-0.104} = -3.57$$

Northern Ireland:

$$\frac{-0.192 + (-0.026 \times 4) + (-0.041)}{-0.117} = -2.88$$

Scotland:

$$\frac{-0.298 + (-0.032 \times 4) + (-0.013)}{-0.091} = -4.87$$

Wales:

$$\frac{-0.165 + (-0.028 \times 4) + (-0.028)}{-0.100} = -3.03$$

The standard errors and 95% CI are calculated using the delta method and are as follows:

| Nation | MRS (absolute) | Standard Error | Lower Confidence Interval | Upper Confidence Interval |
|---|---|---|---|---|
| England | 3.57 | 0.59 | 2.41 | 4.73 |
| Northern Ireland | 2.88 | 0.55 | 1.80 | 3.96 |
| Scotland | 4.87 | 0.93 | 3.04 | 6.70 |
| Wales | 3.03 | 0.59 | 1.88 | 4.18 |

### Estimation of lockdown scenario trade-offs using marginal rates of substitution for excess deaths

The calculation for the marginal rate of substitution (MRS) for introducing a 1-week red restriction lockdown where all routine non-COVID healthcare procedures are postponed is the addition of the MRS for the features that describe the scenario, such that:

$$\frac{\beta_{red\_restrictions}}{-\beta_{excess\_deaths}} + \frac{\beta_{length} \times X_{weeks}}{-\beta_{excess\_deaths}} + \frac{\beta_{all\_health\_postponed}}{-\beta_{excess\_deaths}} \quad (1)$$

This can be simplified as:

$$\frac{\beta_{red\_restrictions} + \beta_{length} \times X_{weeks} + \beta_{all\_health\_postponed}}{-\beta_{excess\_deaths}} \quad (2)$$

Following (2), the MRS for the scenario described above are:

England:

$$\frac{-0.246 + (-0.024 \times 1\ ) + (-0.028)}{-0.118} = -2.53$$

Northern Ireland:

$$\frac{-0.192 + (-0.026 \times 1) + (-0.041)}{-0.133} = -1.95$$

Scotland:

$$\frac{-0.298 + (-0.032 \times 1) + (-0.013)}{-0.146} = -2.35$$

Wales:

$$\frac{-0.165 + (-0.028 \times 1) + (-0.028)}{-0.132} = -1.67$$

Standard errors and 95% Confidence Intervals (CI) are calculated using the delta method and are shown below.

| Nation | MRS (absolute) | Standard Error | Lower Confidence Interval | Upper Confidence Interval |
|---|---|---|---|---|
| England | 2.53 | 0.27 | 2.00 | 3.06 |
| Northern Ireland | 1.95 | 0.28 | 1.39 | 2.50 |
| Scotland | 2.35 | 0.23 | 1.89 | 2.81 |
| Wales | 1.67 | 0.26 | 1.17 | 2.18 |

### Estimation of lockdown scenario trade-offs using marginal rates of substitution for decreases in the ability to buy things

The calculation for the marginal rate of substitution (MRS) for introducing a 2-week red restriction lockdown where all routine non-COVID healthcare procedures are postponed in terms of changes (decreases) in the ability to spend are:

England:

$$\frac{-0.246 + (-0.024 \times 4\ ) + (-0.028)}{-0.104} = -3.57$$

Northern Ireland:

$$\frac{-0.192 + (-0.026 \times 4) + (-0.041)}{-0.117} = -2.88$$

Scotland:

$$\frac{-0.298 + (-0.032 \times 4) + (-0.013)}{-0.091} = -4.87$$

Wales:

$$\frac{-0.165 + (-0.028 \times 4) + (-0.028)}{-0.100} = -3.03$$

The standard errors and 95% CI are calculated using the delta method and are as follows:

| Nation | MRS (absolute) | Standard Error | Lower Confidence Interval | Upper Confidence Interval |
|---|---|---|---|---|
| England | 3.57 | 0.59 | 2.41 | 4.73 |
| Northern Ireland | 2.88 | 0.55 | 1.80 | 3.96 |
| Scotland | 4.87 | 0.93 | 3.04 | 6.70 |
| Wales | 3.03 | 0.59 | 1.88 | 4.18 |
